# Strategies to resolve the gap in Adolescent Tuberculosis care at four health facilities in Uganda: The TEEN TB pilot project

**DOI:** 10.1101/2023.05.25.23290553

**Authors:** Samson Omongot, Winters Muttamba, Irene Najjingo, Joseph Baruch Baluku, Sabrina Kitaka, Stavia Turyahabwe, Bruce Kirenga

## Abstract

**Introduction:** In 2021, an estimated 10.6 million people fell ill with tuberculosis (TB) globally, 1.2 million of these were children. About 40% of them aged between 5 and 14 years with TB are missed annually. In Uganda, 44% of adolescents with chronic cough of ≥2 weeks do not seek care from health facilities. Therefore, strategies to promote health care-seeking behaviour among adolescents are urgently needed. We piloted a project (TEEN TB project) aimed at improving uptake of tuberculosis (TB) care services among adolescents at Ugandan health facilities.

**Methodology:** We developed an adolescent TB awareness and screening package using the human centred design. This technique puts real people at the centre of the development process. The package consisted of 3 interventions (TB screening cards, adolescent-TB awareness poster messages and a local TB awareness song) deployed in the project health facilities and their surrounding communities. Data on socio-demographic and clinical characteristics of adolescents were collected for the period between October 2021 and March 2022 at 4 project health facilities (Kawolo, Iganga, Gombe and Kiwoko). We collected before and after intervention data from facility level records to determine the effect of the package.

**Results:** A total of 394 adolescents were included and the majority (76%) were still in school. Overall, the intervention improved adolescent TB care in the four project health facilities. The average number of adolescents screened increased by 94% from 159 to 309, with an incidence rate ratio (IRR) of 1.9 (95% CI: 1.9-2.0, p <0.001), there was a 2-fold increase among those presumed to have TB; from 13 to 29, IRR of 2.2 (95% CI: 1.9-2.5, p <0.001) and those tested with GeneX-pert and microscopy increased more than 3 times from 8 to 28, IRR of 3.3 (95% CI: 2.8-3.8, p <0.001). There was a minimal increase in the average monthly number of adolescents with a positive result from 1.6 to 2.4 and linkage to TB care services from 2 to 3.1. These were not statistically significant at p=0.170 and p=0.154 respectively.

**Conclusion:** The project improved uptake of TB services among adolescents along the TB care cascade (screening, TB testing and linkage to care). We recommend a robust and fully powered randomized controlled trial to evaluate the effectiveness of the package.

## Introduction

Globally, an estimated 10.6 million people fell ill with tuberculosis (TB) in 2021; among these 1.2 million were children ^1^. Previous, in 2020 the incidence of TB was 220 TB cases per 100, 000 populations in Africa^2^. In the same year, a total case notification of 1.4 million with TB treatment coverage of 56%, and a TB case fatality ratio of 22% was reported ^2^. In Uganda, TB incidence stands at 196 cases per 100,000 populations, treatment coverage at 68%, case fatality ratio at 19%, with 62,526 cases notified ^2^. Globally, 36% of TB cases are missed in health centres, and these are either undiagnosed or unreported ^3^. The prevalence to notification ratio stands at 1.7. Globally, about 40% of TB cases among children aged between 5 and 14 years are missed annually ^4^. There is currently no routine reporting of TB data among adolescents (10-19 years). Additionally, there is paucity of epidemiological data to guide interventions that address the specific needs of adolescents ^5^ . One of the barriers to TB case finding in adolescents is low rates of TB screening and testing in this age group, as demonstrated in the Uganda National TB survey which revealed that only 56% of adolescents reporting a chronic cough of ≥2 weeks sought care from health facilities. Only 3% of these were asked to provide sputum and 1.8% were asked to undergo a chest X-ray (CXR) examination ^6^. Although there are several other reasons for the low TB testing in adolescents, lack of skills and knowledge by the health care workers to deliver adolescent friendly health services are some of the leading barriers ^7^ . Studies have revealed that delays in diagnosis, stigma related with diagnosis and treatment, long waiting hours at health facilities, absence of nutritional support for patients with TB, and absence of comprehensive psychosocial support programs are barriers to access and adherence to TB care ^7, 8^. Strategies including community engagement, training health workers and strengthening public-private partnerships have been found vital in TB control and reducing the missed cases ^9^. Strategies to promote care-seeking behaviours among adolescents (demand creation) and ensuring adequate evaluation in health facilities are needed to bridge the gap in the quality of TB care among adolescents. In response to this, we piloted an adolescent TB care package at four health facilities aimed at improving adolescent TB care seeking behaviour.

## Methods

### Project design

A human centred design was used to develop the adolescent-friendly TB awareness and screening package. The package comprised of adolescent TB educative and informative messages simulated in form of educational posters, TB awareness local song (“*Bulamu bwo*”) and TB screening cards. This package was implemented in four (4) project health facilities. Implementation phase had a number of activities that were kick started by a meeting with relevant stakeholders (Ministry of Health, educational and political leaders, and key adolescent health care providers). The meeting was led by the National TB programme with the research team providing technical support. During this meeting, the project team presented data on the clinical and economic burden and unmet needs of TB among adolescents. Besides engagement of different stakeholders, health workers were given three (3) days training by TB programme staff, assisted by the research team. The training focused on Four (4) key ingredients of adolescent friendly health services such as; being non-judgmental, friendly adolescent services, service demand creation and community awareness/ support. Implementation of TB awareness package was done through the support of project volunteers, research assistants, village health teams (VHTs) and health workers. The VHTs displayed posters on collection points of adolescents in the communities, in the project health facilities and drug shops/stores to sensitize adolescents to seek TB care. The VHTs also identified and engaged local radios serving respective communities to play the local song. The health workers and volunteers oriented the adults who sought care in the TB clinics or with TB symptoms and had adolescents at home on filling of the TB-screening cards. These adults were eventually given the cards to screen their adolescents at home, they were asked to send adolescents found with TB symptoms to the project health facilities to seek care. Poster messages and screening cards were implemented for six (6) months (October, 2021 to March, 2022). The local song was played on the radio stations for 3 months (January to March 2022). Adolescents who reported to the health facility from October 2021 to March 2022 were subjected to TB screening; those who presented with any of the four symptoms (cough, fever, drenching night sweats and weight loss) were consented (≥ 18years), or assented (< 18 years) and enrolled into the project. These adolescents were taken through the entire TB care cascade (screening, testing and linkage). All responses on socio-demographic and clinical characteristics of the enrolled adolescents were captured into a computer tablet. This data was retrieved and analyzed. To assess the impact of the package, before and after intervention data for adolescents screened, presumptive, GeneXpert tested, positive for TB and those linked to TB care was collected from the health facility registers and analyzed.

### Project settings

The study sites included health centers and district level hospitals that were randomly selected. These were health facilities with TB diagnostic and treatment units located in both rural and urban places in the central region of Uganda. Among the four health facilities selected, Kiwoko and Gombe hospitals were rural, while Iganga and Kawolo hospitals were urban. Gombe is a public health facility with 100 bed capacity, while Kiwoko is a faith-based private health facility with 204 bed capacity. The other two facilities (Iganga and Kawolo hospitals) are both public urban health facilities with up to 100 bed capacity. All the health facilities offer TB and other services, such as out-patients, in-patients, antenatal, HIV, eye, dental, nutrition and community services.

### Project participants

The project participants included adolescents who presented to the health facilities at different service delivery points such as outpatient department, HIV/ART clinic, maternal and reproductive health clinic (MRH), inpatient wards and TB clinics. We included all adolescents aged 10-19 years with at least one symptom suggestive of TB; predefined according to the World Health Organisation’s (WHO) criteria (cough for ≥2 weeks; persistent fever for ≥2 weeks; noticeable weight loss; and excessive night sweats). TB patients who were already on TB treatment or had been screened within two weeks were excluded.

### Project procedures

#### Data collection

Adolescents (10-19 years) who came to the project health facilities were identified from the service delivery points (OPD, ART clinic, MRH clinic, TB clinic and wards) and screened for TB using the national screening algorithm. Those with at least one of the TB symptoms and not on anti-TB treatment were considered eligible and were consented (≥ 18years) or assented (≤ 18 years) by their parents or health workers and enrolled into the project. Identification numbers were provided and electronic interviewer-administered structured questionnaires were used to collect data from each of the adolescents who participated in the project. All the enrolled adolescents were subjected to TB testing using either GeneXpert or smear microscopy. Those who tested TB positive were initiated on treatment, while the negative ones were treated as per the TB national guidelines.

#### Data management/analysis

Socio-demographics and clinical characteristics data were collected from the adolescents. Aggregate level data was collected from health facility registers for different variables such as number screened, number presumed and number tested. This data was collected 6 months 2016 and analyzed with STATA version 14 software. Descriptive univariate analysis was conducted to get the averages per facility, standard deviation plus lower and upper ranges. Bivariate analysis was used to determine statistical associations between variables. Pearson chi-square test was used to compare association in categorical variables; statistically significant associations were rated at p-value of less than 0.05 at 95% confidence interval (CI). T-test was conducted to test significance of difference in means before and after intervention. Logistic Regression analysis was done to get Incidence Rate Ratios (IRR). Graphs were plotted using STATA software. Socio-demographics and clinical characteristics were descriptively summarized in tabular form as frequencies and percentages. Before and after intervention data analysis was presented inform of bar graphs and box plots with interquartile ranges.

#### Ethics approvals and informed consent

Ethical approvals were obtained from the Mulago Hospital Research Ethics Committee (MHREC 1922) and the Uganda National Council of Science and Technology (UNCST HS1042ES). Administrative approval (ADM. 105/309/05) was obtained from Ministry of health. All participants gave written informed consents, while adult caregivers and health workers gave written informed assents.

## Results

### Socio-demographic characteristics

Data on socio-demographics was collected during the intervention phase to assess the response to the implemented package in-terms of the different characteristics.

Three hundred ninety four (394) adolescents were enrolled. Majority, 198 (50%) were aged between 10-15 years, they were mainly females 255(65%), and still in school 298(76%). Majority, 336(85%) were unmarried. Kawolo hospital enrolled the highest number, 137(35%) of adolescents compared to the other health facilities as shown in table 1 bellow.

**Table 1.** - Socio-demographic characteristics.

| Characteristics | Items | Frequency | Percentage (%) |
| --- | --- | --- | --- |
| Age | 10- 15 years | 198 | 50.3 |
|  | 16- 20 years | 196 | 49.7 |
| Sex | Male | 139 | 35.3 |
|  | Female | 255 | 64.7 |
| Employment status | Student | 298 | 75.6 |
|  | Employed | 30 | 7.6 |
|  | Unemployed | 66 | 16.8 |
| Marital status | Married | 58 | 14.7 |
|  | Single | 336 | 85.3 |
| Educational Level | Complete primary | 23 | 5.8 |
|  | Incomplete primary | 176 | 44.7 |
|  | Complete secondary | 28 | 7.1 |
|  | Incomplete secondary | 145 | 36.8 |
|  | Tertiary | 5 | 1.3 |
|  | Never attended school | 17 | 4.3 |
| Site | Kawolo Hospital | 137 | 34.8 |
|  | Iganga Hospital | 88 | 22.3 |
|  | Kiwoko Hospital | 77 | 19.5 |
|  | Gombe Hospital | 92 | 23.4 |

### Clinical characteristics

Data on clinical characteristics was collected to assess for additional information on TB awareness by the adolescents as described below.

Up to 330 (84%) of the adolescents had ever heard about TB, and majority; 322(82%) reported no history of TB contact. Up to 49 (68%) of the adolescents reported to have had TB contact with family members. Most of participants; 265(67%) knew their HIV status, 60(15%) of these reported HIV positive status, and 59 (98%) of them were on ART. With regard to risk factors, majority 387(98%) had never smoked, and 357(91%) did not consume alcohol. Majority, 326(83%) had heard about adolescent TB, of which, 266(38%) reported to have got information from the research team that was in the facility. All this information is shown on table 2 below.

**Table 2.** - Clinical characteristics.

| Characteristic | Item | Frequency | Percentage |
| --- | --- | --- | --- |
| Do you know about TB? | Yes | 330 | 83.8 |
|  | No | 64 | 16.2 |
| Close contact with someone with TB or chronic cough? | Yes | 72 | 18.3 |
|  | No | 322 | 81.7 |
| If yes, relationship with contact? | Family members | 49 | 68.1 |
|  | Family friend | 11 | 15.3 |
|  | Fellow students | 12 | 16.7 |
| Do you know your HIV status? | Yes positive | 60 | 15.2 |
|  | Yes negative | 205 | 52.0 |
|  | I don't know | 129 | 32.7 |
| If yes and positive, are you on ART? | Yes | 59 | 98.3 |
|  | No | 1 | 1.7 |
| Do you smoke tobacco? | Yes | 7 | 1.8 |
|  | No | 387 | 98.2 |
| Do you take alcohol? | Yes | 37 | 9.4 |
|  | No | 357 | 90.6 |
| Have you taken an x-ray? | Yes | 57 | 14.5 |
|  | No | 337 | 85.5 |
| Have you collected sputum? | Yes | 305 | 77.4 |
|  | No | 89 | 22.6 |
| Have you heard about adolescent TB? | Yes | 326 | 82.7 |
|  | No | 68 | 17.3 |
| If yes, source of information about adolescent TB? | TEEN- TB Team | 266 | 37.6 |
|  | Radio/TV | 138 | 19.5 |
|  | Friend(s) | 83 | 11.7 |
|  | VHTs | 67 | 9.5 |
|  | Local song | 65 | 9.2 |
|  | Social media | 46 | 6.5 |
|  | Posters | 33 | 4.7 |
|  | Screening cards | 9 | 1.3 |

### Impact of package on TB care cascade

The impact of the package on adolescent TB care is presented along the TB care cascade In Figure 1, 2, Tables 3, 4 and 5.

**Figure 1.**
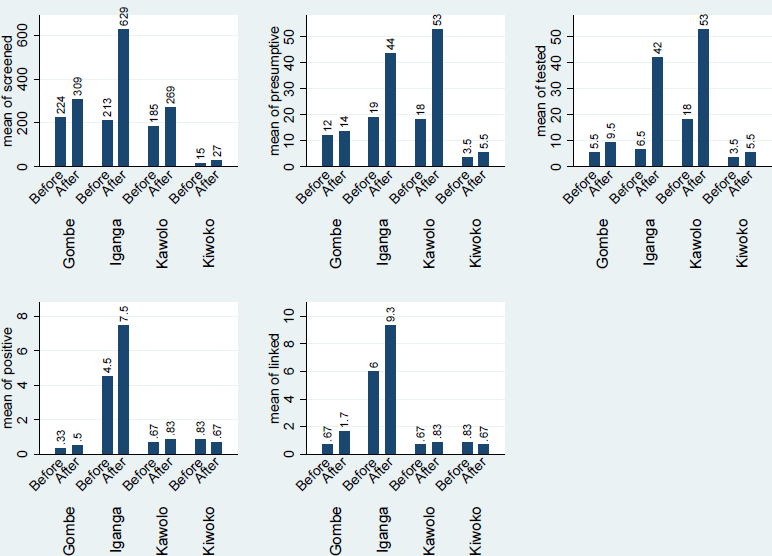
Bar graphs showing the before & after intervention average monthly scores of adolescents screened, presumptive, tested, positive and linked to care at each of the project health facilities.

**Table 3:** Incidence rate ratio (IRR) of adolescents screened for TB.

|  |  | **Mean\*** |  | **Likelihood** |  |  |
| --- | --- | --- | --- | --- | --- | --- |
| **Facility** | **Period** | **Mean(#screened)** | **p-value** | **IRR** | **p-value** | **95% CI** |
| **Overall** | Before | 159 |  |  |  |  |
|  | After | 309 | 0.025 | **1.9** | <0.001 | (1.9-2.0) |
| Gombe Hospital | Before | 224 |  |  |  |  |
|  | After | 309 | 0.190 | 1.4 | <0.001 | (1.3-1.5) |
| Iganga Hospital | Before | 213 |  |  |  |  |
|  | After | 629 | 0.040 | 3.0 | <0.001 | (2.8-3.2) |
| Kawolo Hospital | Before | 185 |  |  |  |  |
|  | After | 269 | 0.123 | 1.5 | <0.001 | (1.4-1.6) |
| Kiwoko Hospital | Before | 15 |  |  |  |  |
|  | After | 27 | 0.071 | 1.8 | <0.001 | (1.4-2.4) |

\* t-Test

**Table 4:** Incidence rate ratio (IRR) for TB presumptive adolescents

|  |  | **Mean\*** |  | **Likelihood** |  |  |
| --- | --- | --- | --- | --- | --- | --- |
| **Facility** | **Period** | **Mean(# presumptive)** | **p-value** | **IRR** | **p-value** | **95% CI** |
| **Overall** | Before | 13 |  |  |  |  |
|  | After | 29 | 0.019 | **2.2** | <0.001 | (1.9-2.5) |
| Gombe Hospital | Before | 12 |  |  |  |  |
|  | After | 14 | 0.356 | 1.1 | 0.467 | (0.8-1.6) |
| Iganga Hospital | Before | 19 |  |  |  |  |
|  | After | 44 | 0.147 | 2.3 | <0.001 | (1.9-2.9) |
| Kawolo Hospital | Before | 18 |  |  |  |  |
|  | After | 53 | 0.005 | 2.9 | <0.001 | (2.3-3.6) |
| Kiwoko Hospital | Before | 4 |  |  |  |  |
|  | After | 6 | 0.223 | 1.6 | 0.105 | (0.9-5.4) |

\*t-Test

**Table 5:** Incident rate ratio (IRR) of adolescents tested for TB

|  |  | **Mean\*** |  | **Likelihood** |  |  |
| --- | --- | --- | --- | --- | --- | --- |
| **Facility** | **Period** | **Mean(# Tested)** | **p-value** | **IRR** | **p-value** | **95% CI** |
| **Overall** | **Before** | 8 |  |  |  |  |
|  | **After** | 28 | 0.007 | **3.3** | <0.001 | (2.8-3.8) |
| **Gombe** | **Before** | 6 |  |  |  |  |
|  | **After** | 10 | 0.102 | 1.7 | 0.012 | (1.1-2.7) |
| **Iganga** | **Before** | 7 |  |  |  |  |
|  | **After** | 42 | 0.072 | 6.5 | <0.001 | (4.6-9.1) |
| **Kawolo** | **Before** | 18 |  |  |  |  |
|  | **After** | 53 | 0.005 | 2.9 | <0.001 | (2.3-3.6) |
| **Kiwoko** | **Before** | 4 |  |  |  |  |
|  | **After** | 6 | 0.223 | 1.6 | 0.105 | (0.9-2.7) |

\*t-Test

The entire analysis revealed the following results:

### Screening for TB

There was an increase in the numbers of adolescents screened for TB after the intervention across all the four project health facilities. The overall average increase was 94% (159 to 309). This increase resulted in 1.9 incident rate ratio (IRR) at 95% CI: 1.9-2.0 and this was statistically significant (p <0.001). Iganga hospital had a threefold increase in the number screened from 213 to 629. The increase gave rise to 3.0 IRR at 95% CI: 2.8-3.2 and was statistically significant (p < 0.001). Kawolo hospital had 45% (185 to 269) resulting in 1.5 IRR at 95% CI: 1.4-1.6. This increase was statistically significant (p < 0.001). Kiwoko Hospital had 80% (15-27) resulting in 1.8 IRR at 95% CI: 1.4-2.4 the increase was statistically significant (p <0.001) and Gombe hospital 38% (224 to 309), giving rise to 1.4 IRR at 95% CI: 1.3-1.5, and was statistically significant (p <0.001). The details of numbers screened before and after the intervention are indicated in Figures 1 and 2 and Table 3.

**Figure 2.**
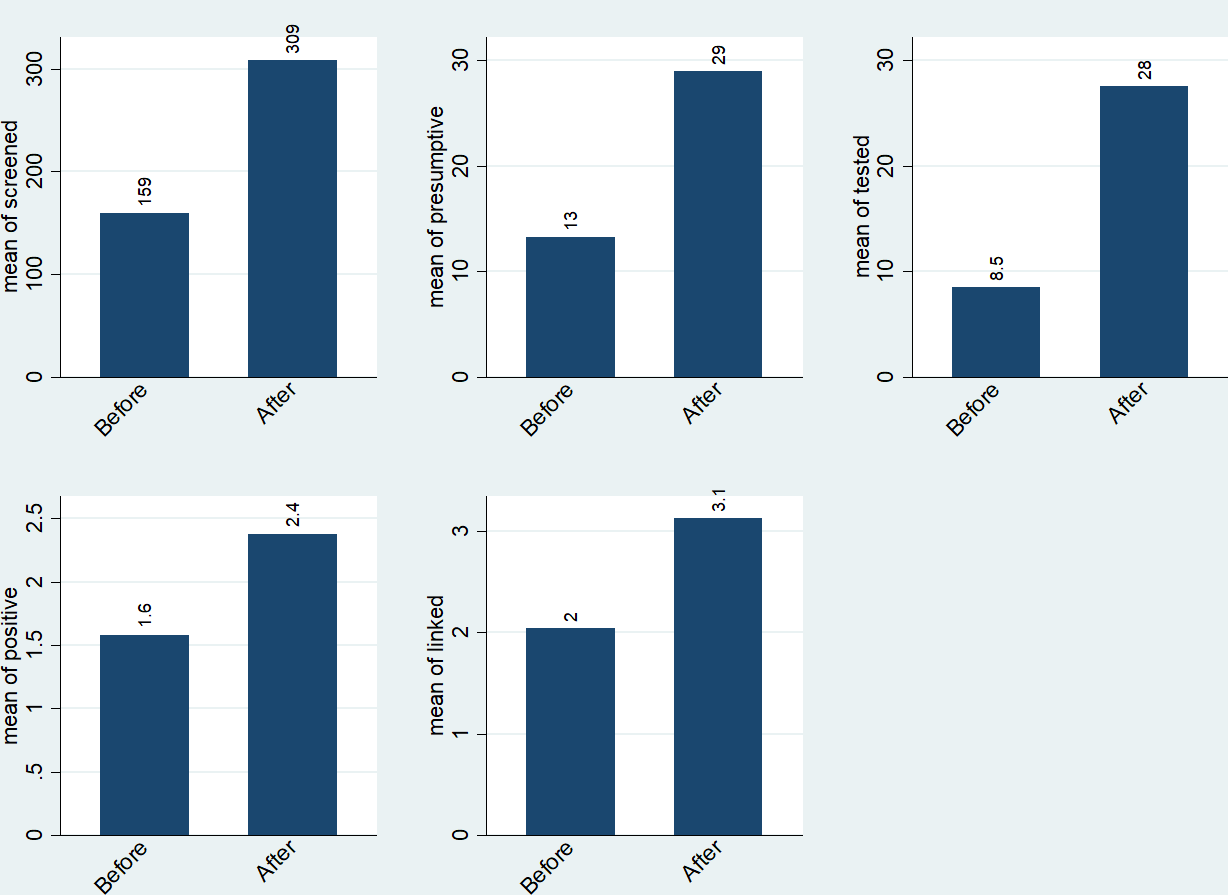
Bar graph showing the overall six months average scores for adolescents screened, presumptive, tested, positive and linked to treatment and care before & after the intervention.

### TB Presumptive patients

There was an increase in the average number of presumptive TB adolescents identified during the project period. The overall average increase after the intervention was from 13 to 29, thus, more than double the number before. This resulted in 2.2 incident rate ration (IRR) at 95% CI, 1.9-2.5 that was statistically significant (p <0.001). Iganga hospital had more than double increase from 19 to 44. The increase gave rise to an IRR of 2.3 at 95% CI: 1.9 −2.9, and this was statistically significant (p < 0.001). Kawolo hospital had a threefold increase from 18 to 53 resulting in an IRR of 2.9 at 95% CI: 2.3 −3.6, this was statistically significant (p < 0.001). Kiwoko hospital had 50% (4 to 6) resulting in an IRR of 1.6 at 95% CI: 0.9-5.4, an increase that was not statistically significant (p= 0.105) and Gombe hospital had 17% (12 to 14), giving rise to an IRR of 1.1 at 95% CI: 0.8-1.6, this was not statistically significant (p=0.467). The details of the presumptive TB before and after the intervention are indicated in Figures 1 and 2 and Table 4.

### TB testing

The overall average number of adolescents tested for TB increased from 8 to 28 after the intervention, this was more than threefold increase resulting in an IRR of 3.3 at 95% CI: 2.8-3.8. This increase was statistically significant (p <0.001). The increase was more than six - fold in Iganga hospital from 7 to 42 resulting in an IRR of 6.5 at 95% CI: 4.6-9.1 which was statistically significant (p <0.001). Kawolo hospital had a threefold increase from 18 to 53 resulting in an IRR of 2.9 at 95% CI: 2.3-3.6, which was statistically significant (p <0.001). Gombe hospital had 67% (6 to 10), resulting in an IRR of 1.7 at 95% CI: 1.1-2.7, which was statistically significant (p=0.012). Kiwoko hospital was at 50% (4 to 6), resulting in an IRR of 1.6 at 95% CI: 0.9-2.7, although it was not statistically significant (p=0.105). The numbers tested for TB are indicated in Figure 1 and 2 and Table 5.

### TB Positive adolescents

Overall, there was minimal average increase in the number of TB positive adolescents identified after the intervention by 50% (2 to 3), and this was not statistically significant (p=0.170). An increase of 60% (5 to 8) was registered in Iganga hospital, 25% (4 to 5) in Gombe hospital while Kawolo hospital had an increase of 24% (67 to 83). Kiwoko hospital registered a reduction by-16% (0.83 to 0.67) as indicated in Figures 1 and 2.

### Linkage to TB care and treatment

There was minimal improvement in linkage of adolescents to treatment and care, there was 50% (2 to 3.1) increase in the number linked after the intervention, though not statistically significant (p=0.154). Gombe hospital had more than two-fold increase from 67 to 170, Iganga hospital had 50% increase (6 to 9), Kawolo hospital had an increase of 24% (67 to 83). Kiwoko hospital registered a decline at −16% (83 to 67) as shown in Figures 1 and 2.

## Discussion

The project sought to resolve the gap in adolescent TB care seeking behavior using a human centered design approach. An adolescent TB awareness and screening package was developed and piloted in four health facilities in central Uganda. Generally findings indicated a statistically significant increase in the number of adolescents screened, TB presumptive and those tested for TB. This significant increase was a result of broadening strategies to disseminate TB information to sensitize adolescents to seek care. One of the studies on adolescent TB had a similar argument by suggesting a comprehensive response to adolescent TB 10

Female adolescents participated more in this study as compared to males, a trend that has been demonstrated by many other adolescent studies, with the high number of females registered being associated with their high susceptibility to TB as compared to their male counterparts ^11.^.

Majority of the TB positives were students. This similar finding was realised in a study that was done in Swaziland schools that captured many TB positive cases from students at schools ^12.^ .Many presumptive cases reported family contacts. Close contact mixing is a common phenomenon demonstrated by a number of studies, for instance the age-and sex-specific social contact study done in the Zambian and South African communities that revealed more than 50% of infections in children resulting from contacts with adult men ^13^. Majority of the enrolled adolescent had no history of smoking or alcohol consumption. Cigarette smoking and alcohol consumption in this case were considered as minimal risk factors for adolescent TB, contrary to this finding, however, some studies have shown close association between TB and cigarette smoking/alcohol consumption ^14^.

While sputum was the main sample used for TB detection as per WHO recommendations on adolescent TB testing, a few x-rays were as well done. These findings related to the 2015 Ugandan TB survey where 1.8% of the participants had X-rays done and 3% of participants asked to provide sputum samples ^6, 15^

Among the key TB awareness interventional approaches that were implemented to close the gap in adolescent TB care seeking, the local song (“Bulamu bwo”) played on radio stations sensitized more adolescents to seek care as compared to TB adolescent poster messages and TB screening done at house hold level by parents or caregivers. Just like this project utilized an awareness strategy, a study in Bangladesh similarly emphasised on increased awareness and service delivery by health care workers as promoters and motivators of increased health care seeking behaviour ^16^

Besides the three key intervention approaches, many received information about TB from the research teams in the health facilities, community village health teams (VHTs) and social media. Apart from lack of information, other similar studies have also documented stigma, delays in diagnosis, long waiting hours, absence of nutritional support and psychosocial support as related factors associated with poor TB health care seeking ^8, 17^

Screening done by parents or guardians at household level demonstrated a potential community approach to reach and mobilize adolescents to seek care. This kind of intervention is a way to enhance contact tracing, which many studies have documented as an approach to easily reach and mobilize TB presumptive patients for testing, ^18^

Most of the adolescents that were enrolled into the project had prior knowledge on TB and had ever heard about adolescent TB, this was an encouraging finding given that other studies have shown lack of knowledge as a leading barrier to TB health care seeking ^19^

Before and after project implementation data obtained from the health facility registers demonstrated a positive impact on numbers screened, presumptives and those tested using GeneXpert. However, this project registered minimal numbers of adolescents who tested positive for TB and linked to care. Despite the minimal numbers, more TB positives were registered in urban project health facilities that had more reliable stations that played the local song as compared to the rural health facilities. This kind of finding was similarly shown in a study done on the burden of adolescent TB in rural eastern Uganda in which a low incident rate was registered ^20^

## Conclusion

The intervention generally improved adolescent TB care seeking behavior along all the important steps of the TB care cascade (screening, TB testing, TB positives and linkage to care). The use of human centered design that directly involved the adolescents in preparing the interventions was a real strength for the project success. However, being a pilot project implemented at the health facilities, we were not able to reach out to other adolescent communities such as schools and distant villages. These results are unlikely to be generalizable. Secondly, our study depended on self-reported responses, which could be affected by recall bias. Out of the many intervention methods that were provided by the expert designers, we were able to only implement three (3) of them due to COVID-19 restrictions, hence impacting on the number of adolescents reached. Additionally, it was not possible to come up with appropriate comparisons of the three interventions because they were not implemented at the same time, although conclusions were drawn after harmonizing the intervention periods.

Given the above said limitations, tthere is a great need to devise mechanisms to mobilize adolescents to seek TB care. Since this was a pilot project, we recommend a robust and fully powered randomized controlled trial to mobilize more adolescents and more so evaluate the effectiveness of the package.

## Data Availability

Most of the Data is in the manuscript, however additional information can be provided when requested

## Acknowledgements

The entire implementation process and success of TEEN-TB project is attributed to the great contribution of different stakeholders, we specifically thank the national TB and leprosy program administration (MOH) who provided administrative clearance and technical support at national level. Design without Borders (DwB) Africa limited team, a creative design group who developed the adolescent TB screening cards & poster messages that were used for household screening and adolescent TB awareness creation respectively. We as well extend our sincere appreciation to the “Ghetto Yute” youth music group who produced the local song (“Bulamu bwo”) with adolescent TB awareness messages that were broadcasted in selected local radio stations within the surrounding communities of the project health facilities. Additionally, we would want to thank the health workers who worked closely with research assistants, village health teams (VHTs) and project volunteers to collect data in the health facilities. Last but not least, Makerere University research and innovation fund for the monetary support that was used to finance all project activities to the end.

